# The impact of the COVID-19 pandemic on wellbeing and cognitive functioning of older adults

**DOI:** 10.1101/2020.08.27.20183129

**Authors:** Sarah De Pue, Céline Gillebert, Eva Dierckx, Marie-Anne Vanderhasselt, Rudi De Raedt, Eva Van den Bussche

**Affiliations:** Brain & Cognition, KU Leuven, Leuven, Belgium; Personality and Psychopathology, Vrije Universiteit Brussel, Brussels, Belgium; Psychiatric Hospital, Alexianen Zorggroep Tienen, Tienen, Belgium; Department of Head and Skin, Ghent University, University Hospital Ghent (UZ Ghent), Ghent, Belgium; Department of Experimental Clinical and Health Psychology, Ghent University, Ghent, Belgium

**Keywords:** COVID-19, aging, wellbeing, cognition

## Abstract

COVID-19 took a heavy toll on older adults. In Belgium, by the end of August, 93% of deaths due to COVID-19 were aged 65 or older. Similar trends were observed in other countries. As a consequence, older adults were identified as a group at risk, and strict governmental restrictions were imposed on them. This has caused concerns about their mental health. Using an online survey, this study established the impact of the COVID-19 pandemic on adults aged 65 years or older, and which factors moderate this impact. Participants reported a significant decrease in activity level, sleep quality and wellbeing during the COVID-19 pandemic. Depression was strongly related to reported declines in activity level, sleep quality, wellbeing and cognitive functioning. Our study shows that the COVID-19 pandemic had a severe impact on the mental health of older adults. This implies that this group at risk requires attention of governments and healthcare.

---

The COVID-19 pandemic took a heavy toll on older adults. For example in Belgium, approximately 41% of the cases were aged 60 or older, but 93% of deaths due to COVID-19 were aged 65 or older (Sciensano, 2020). Similar trends were observed in other countries (e.g., Guterres, 2020). This implied that older adults became a specific focus of several governmental COVID-19 regulations. For example, in Belgium they were no longer allowed to look after their grandchildren, visitors and external services (e.g., meal delivery, cleaning) were not allowed in assisted living facilities and nursing homes for a long time, and people were regularly warned when measures were introduced or relaxed to pay specific attention to older and vulnerable people.

Hence, during the COVID-19 pandemic, older adults were considered as a group at risk. As a response, concerns have risen about the mental health of older adults. The World Health Organization (WHO) has warned that the impact on mental and psychosocial wellbeing of vulnerable groups, such as older adults, will be large and enduring (WHO, 2020). The United Nations (UN) stressed that, although COVID-19 is in the first place a physical health crisis, it has the seeds of a major mental health crisis as well, especially for specific populations such as older adults, if action is not taken (UN, 2020). It has been suggested that the measures taken by governance regarding social distancing and isolation, especially targeting groups at risk, can result in social isolation and loneliness (Banerjee, 2020; Vahia, 2020). The latter variables are known to decrease *wellbeing* and increase the risk for depression and cognitive dysfunction (Brooke & Jackson, 2020). Brooke & Jackson (2020) pointed out that a decline in *activity* and mobility in older adults during lockdown can also lead to more frailty and a lower wellbeing in older adults. Moreover, in response to stress, *sleep quality* can decline and increase the risk for depression (Stanton et al., 2020). Finally, as older adults already face cognitive decline as part of normal aging (Bherer, Erickson, & Liu-Ambrose, 2013; Murman, 2015), which is moderated by several lifestyle variables such as physical activity, engagement in stimulating activities and social network (e.g., Clare et al., 2017), the COVID-19 pandemic causing social isolation and loss of activity, might also impact *cognitive functioning*.

These concerns warrant a thorough assessment of how older adults are currently doing. However, little is hitherto known about the impact of the COVID-19 pandemic on the general population of older adults. A few first studies are being published now, and seem to indicate that older adults’ wellbeing was not severely impacted by the COVID-19 pandemic. However, as the authors indicated, these studies were either conducted in the very early stages of the pandemic when social distancing was recommended by the government but no lockdown was yet installed (Kivi et al., in press; Lopez et al., in press) or in countries such as the Netherlands where at the time of the study the lockdown was not restrictive (van Tilburg et al., in press). Findings of disaster studies might be relevant in this respect. For example, studies on the effect of natural disasters on older adults showed a disaster-related decline in cognitive function (Hikichi et al., 2016). Results on the effect on wellbeing are mixed, and indicated that being older did not necessarily increase psychological vulnerability (Rafiey et al., 2016; see also the “wellbeing paradox”; Hansen & Slagsvold, 2012). However, socialising and social participation were found to be crucial to reduce the risk of cognitive decline and decrease in wellbeing after such disasters (Hikichi et al., 2017), which is precisely what has been severely limited during the COVID-19 pandemic, especially for risk groups such as older adults.

Given these concerns and the lack of studies focused on older adults during the peak of the COVID-19 pandemic, the goal of this study was to establish how adults aged 65 years or older are responding to the COVID-19 pandemic. Using self-report measures in an online survey, the impact of the COVID-19 period on wellbeing, activity level, sleep quality and cognitive functioning was studied. Based on the raised concerns and previous studies on the impact of disasters on older adults, we hypothesised that the COVID-19 pandemic had a detrimental effect on the wellbeing, activity level, sleep quality and cognitive functioning of older adults. Furthermore, we examined possible vulnerability and protective factors that might have influenced the impact of the COVID-19 period on the wellbeing and cognitive functioning of older adults.

## RESULTS

### Participant characteristics and Pearson correlations

Supplementary Table 1 provides a more detailed overview of the participant characteristics. Participants were on average 73 years old (SD=6.99, range 65-97). Figure 1 displays the frequency distribution of age. Two hundred sixty-two were male, 377 were female, and one identified themselves with another gender. Geographically, participants were spread across Flanders (i.e., the northern, Dutch-speaking part of Belgium), as can be seen on Figure 2. The majority of the participants (81%) lived in their own home and 19% lived in an assisted living facility or nursing home (jointly referred to as care facility). 28% lived alone, 54% with one cohabitant and 18% with two or more cohabitants. Most participants (respectively 29%, 41%, 32% and 25%) reported that during the past week they had 3 to 4 contacts (not taking into account cohabitants) in real life outside, no contact in real life inside, 3 to 4 contacts by telephone and more than 9 contacts via the internet (e.g., skype, whatsapp). Most participants (52%) had a university or high school degree and received a monthly individual net income between 1001 and 1500 euro (25%), 1501 and 2000 euro (30%) or 2001 and 2500 euro (17%). The majority of participants (88%) did not suffer from Parkinson’s disease, dementia, stroke, diabetes and/or epilepsy. Only 4% reported that they had been contaminated with COVID-19. 15% reported that at least one of their close relatives or friends had been contaminated with COVID-19.

**Figure 1.**
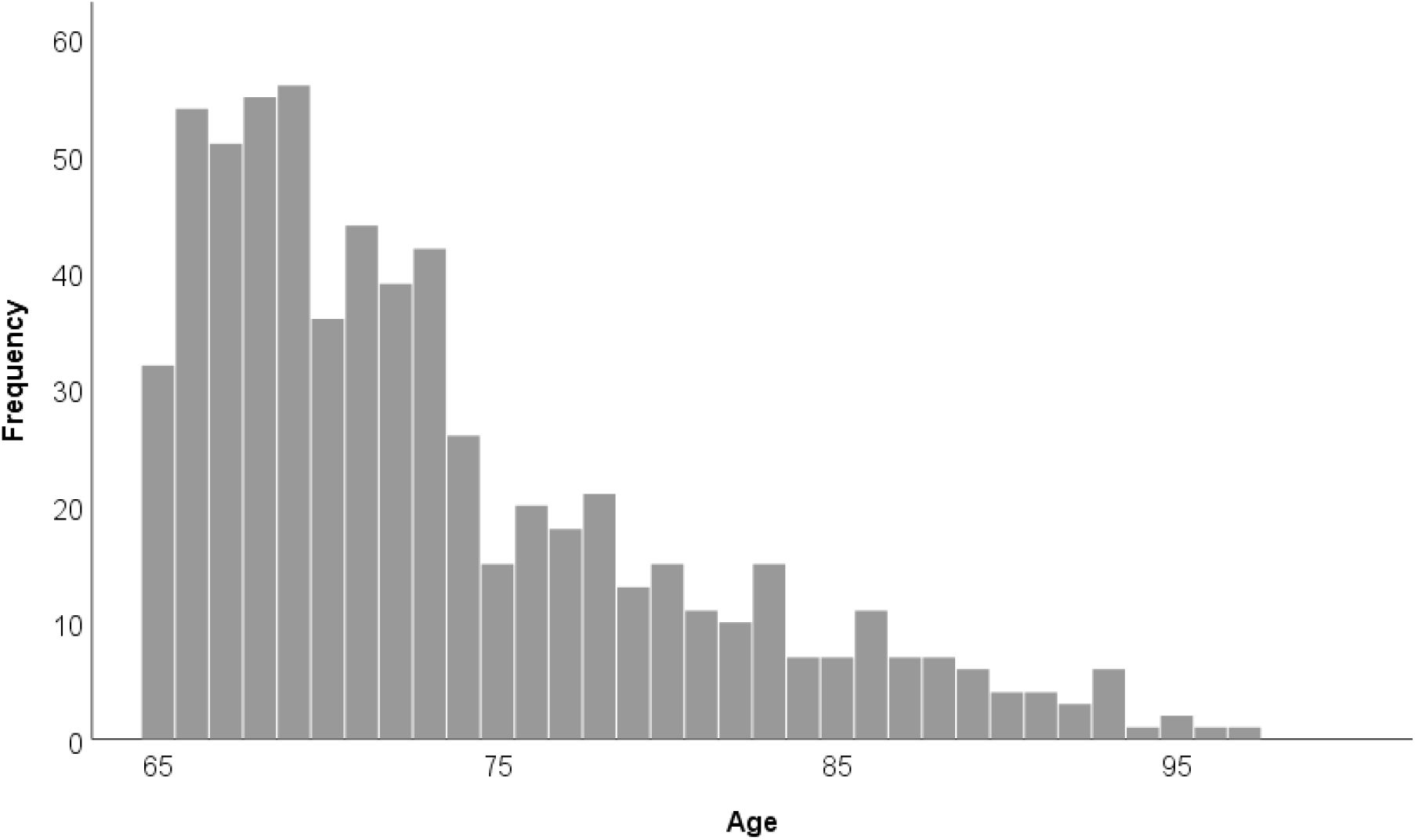
Frequency distribution of age.

**Figure 2.**
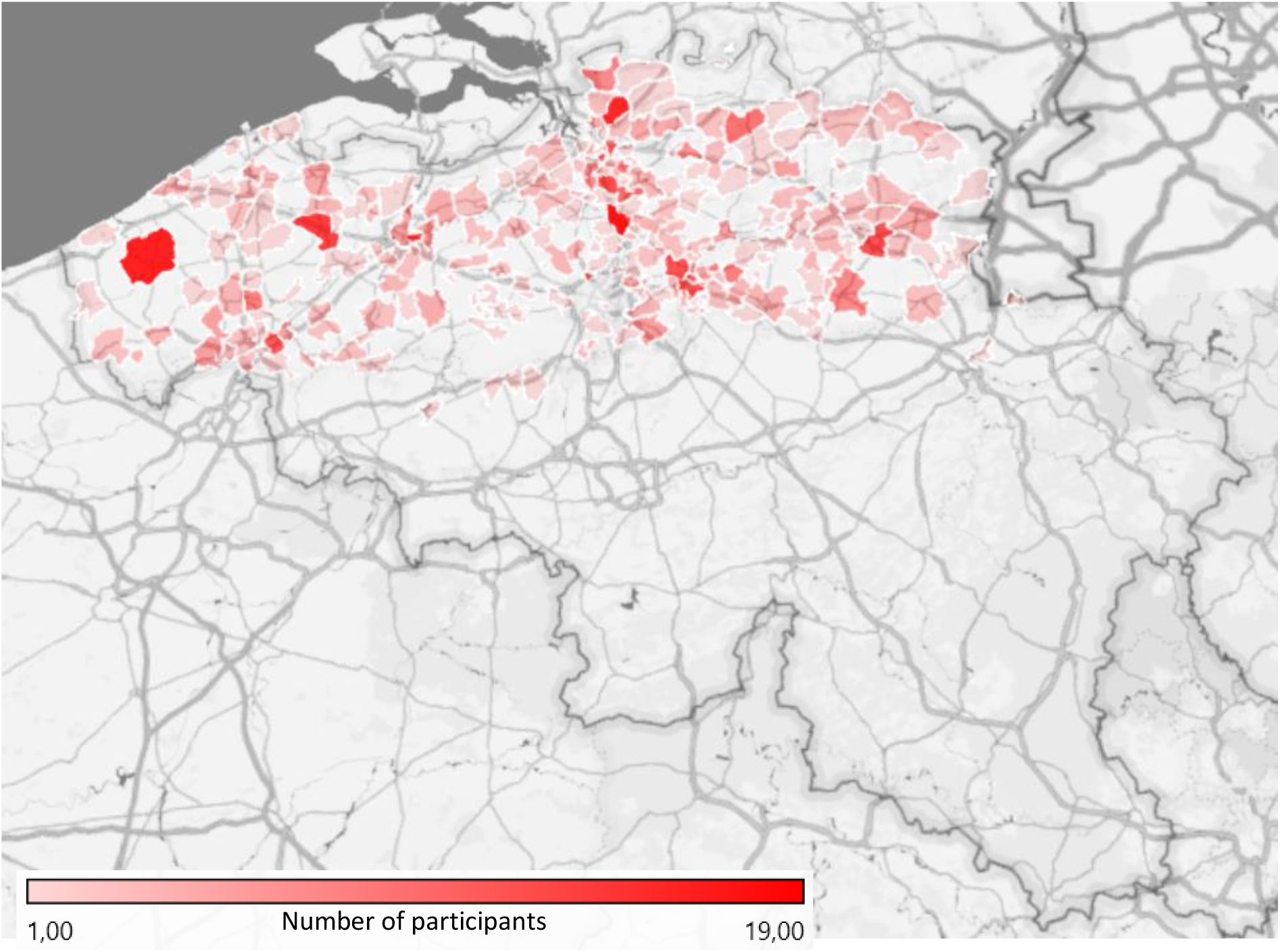
Spreading of participants across Flanders.

The means (SD) of the total scores of the questionnaires and the Pearson correlations between age and the total scores of the questionnaires in the survey can be found in Table 1 (see also Supplementary Figure 1 for a matrix scatterplot containing all two-by-two scatter plots).

### Comparison before and during the COVID-19 period

Table 2 reports the number of participants who reported a decrease, no change or an increase with regards to wellbeing, activity level, sleep quality and cognitive functioning during the COVID-19 period.

**Table 1.** Means (standard deviations) and Pearson correlations between age and the total scores of the questionnaires in the survey.

|  | Valid N | Mean (SD) | Age | CFQ | GDS-15 | PWI-A_pre | PWI-A_current | LSNS-6 | BRS |
| --- | --- | --- | --- | --- | --- | --- | --- | --- | --- |
| Age | 640 | 73 (6.99) | - | -.19*** | -0.005 | -.045 | .020 | -.17*** | -.009 |
| CFQ | 623 | 22.43 (12.02) |  | - | .28*** | -.29*** | -.31*** | -.035 | -.29*** |
| GDS-15 | 640 | 3.00 (3.01) |  |  | - | -.47*** | -.70*** | -.32*** | -.45*** |
| PWI-A_pre | 640 | 78.05 (11.20) |  |  |  | - | .74*** | .33*** | .42*** |
| PWI-A_current | 640 | 70.67 (14.84) |  |  |  |  | - | .35*** | .43*** |
| LSNS-6 | 636 | 16.52 (5.65) |  |  |  |  |  | - | .20*** |
| BRS | 633 | 3.33 (0.67) |  |  |  |  |  |  | - |

*Note.* CFQ = Cognitive Failures Questionnaire total score; GDS-15 = Geriatric Depression Scale-15 total score; PWI-A\_pre = Personal Wellbeing Index – Adults total score retrospectively assessed for the period before COVID-19; PWI-A\_current = Personal Wellbeing Index – Adults total score assessed for the current month (i.e., during the COVID-19 period); LSNS-6 = Lubben Social Network Scale-6 total score; BRS = Brief Resilience Scale mean score; \*\*\* $p < .001$ .

**Table 2.** Mean (SD) of the reported changes in wellbeing, activity level and sleep quality (all based on the calculated difference scores) and cognitive functioning (based on the subjective cognitive change questions) and the number of participants who reported a decrease (N_decrease_), no change (N_nochange_) or an increase (N_increase_) in wellbeing, activity level, sleep quality and cognitive functioning based on the difference scores and the subjective cognitive change questions.

| | | *M(SD)* | $N_{decrease}$ | $N_{nochange}$ | $N_{increase}$ |
| --- | --- | --- | --- | --- | --- |
| Change in wellbeing (PWI-A) | General life satisfaction | -9.63 (15.34) | 317 | 299 | 24 |
|  | Standard of living | -3.98 (11.41) | 140 | 479 | 21 |
|  | Health | -2.88 (9.39) | 130 | 484 | 26 |
|  | Achieving in life | -3.88 (11.09) | 131 | 491 | 18 |
|  | Relationships | -7.09 (15.09) | 213 | 398 | 29 |
|  | Safety | -10.39 (16.10) | 291 | 340 | 9 |
|  | Community connectedness | -10.89 (17.70) | 290 | 323 | 27 |
|  | Future security | -10.39 (14.69) | 306 | 328 | 6 |
|  | Subjective wellbeing (PWI-A total score) | -7.39 (10.02) | 483 | 113 | 44 |
| Change in activity level |  | -0.94 (1.61) | 321 | 276 | 43 |
| Change in sleep quality |  | -0.28 (1.19) | 122 | 472 | 46 |
| Change in cognitive functioning | Cognitive functioning | 1.94 (0.31) | 50 | 576 | 14 |
|  | Remembering | 2.93 (0.40) | 53 | 575 | 12 |
|  | Concentration | 2.91 (0.44) | 78 | 539 | 23 |
|  | Doing two things at the same time | 2.97 (0.35) | 37 | 584 | 19 |
|  | Recalling | 2.92 (0.41) | 63 | 560 | 17 |
|  | forgetfulness | 2.92 (0.40) | 63 | 560 | 17 |

*Note.* For the subjective cognitive change questions assessing problems with remembering, concentration, doing two things at the same time, recalling and forgetfulness, the response options 1="a lot more problems than before" and 2="more problems than before" were grouped together and the response options 4="less problems than before" and 5="a lot less problems than before" were grouped together in the reported numbers of participants.

With regards to wellbeing, the most prominent decreases were reported for general life satisfaction, safety, community connectedness and future security (see Table 2). The subjective wellbeing score indicated that the majority of the participants (76%) reported a decrease in wellbeing in one or more domains. Half of the participants reported that their activity level had decreased and 19% reported a decreased sleep quality during the COVID-19 period. A minority of the participants (8%) reported that their cognitive functioning in general had decreased during the COVID-19 period, with 8%, 12%, 6%, 10% and 10% indicating increases in problems with remembering things, concentrating on something, doing two things at the same time, recalling things or forgetfulness, respectively.

Paired-samples *t*-tests with Bonferroni correction (*α*=.0045) indicated that participants 113 reported that they had been significantly less active during the past week (i.e., during the COVID-19 114 period) compared to before the COVID-19 period (*M*_current_=6.65; *M*_before_=7.58), *t*(639)=-14.69, *p<*.001, 115 *d*=.58. They also reported that their quality of sleep had been poorer during the past week compared 116 to before COVID-19 (*M*_current_=6.88; *M*_before_=7.16), *t*(639)=-5.87, *p<*.001, *d*=.23. Furthermore, life 117 satisfaction in all seven domains, general life satisfaction and subjective wellbeing were significantly 118 lower currently compared to before COVID-19: *t*(639)=8.84, *p<*.001, *d*=.35 for standard of living 119 (*M*_current_=76; *M*_before_=80), *t*(639)=7.75, *p<*.001, *d*=.31 for health (*M*_current_=73; *M*_before_=76), *t*(639)=8.84, 120 *p<*.001, *d*=.35 for achieving in life (*M*_current_=75; *M*_before_=78), *t*(638)=11.88, *p<*.001, *d*=.47 for 121 relationships (*M*_current_=71; *M*_before_=79), *t*(639)=16.33, *p<*.001, *d*=.64 for safety (*M*_current_=70; *M*_before_=81), 122 *t*(639)=15.57, *p<*.001, *d*=.61 for community connectedness (*M*_current_=66; *M*_before_=77), *t*(639)=17.90, 123 *p<*.001, *d*=.71 for future security (*M*_current_=66; *M*_before_=76), *t*(639)=15.88, *p<*.001, *d*=.63 for general life 124 satisfaction (*M*_current_=69; *M*_before_=78) and *t*(639)=18.66, *p<*.001, *d*=.74 for the subjective wellbeing 125 index (*M*_current_=71; *M*_before_=78). Note that when participants filled in the survey (i.e., week 1 to 5; see 126 also Supplementary Table 1) did not lead to significant systematic fluctuations with regards to the 127 reported changes in wellbeing, activity level, sleep quality and cognitive functioning.

### Linear and ordinal logistic regression analyses

Supplementary Tables 2 and 3 report the Pearson correlations between the difference scores and subjective cognitive change questions and potential continuous vulnerability and protective variables, as well as the means (SD) for the difference scores and subjective cognitive change questions for each level of the potential categorical vulnerability and protective variables. Linear and ordinal logistic regression analyses were conducted to study possible vulnerability and protective factors. Table 3 provides the model fit of all models and the statistics for the predictors who made a significant contribution to the models. No problems of multicolinearity arose (all VIF<1.78).

**Table 3.**
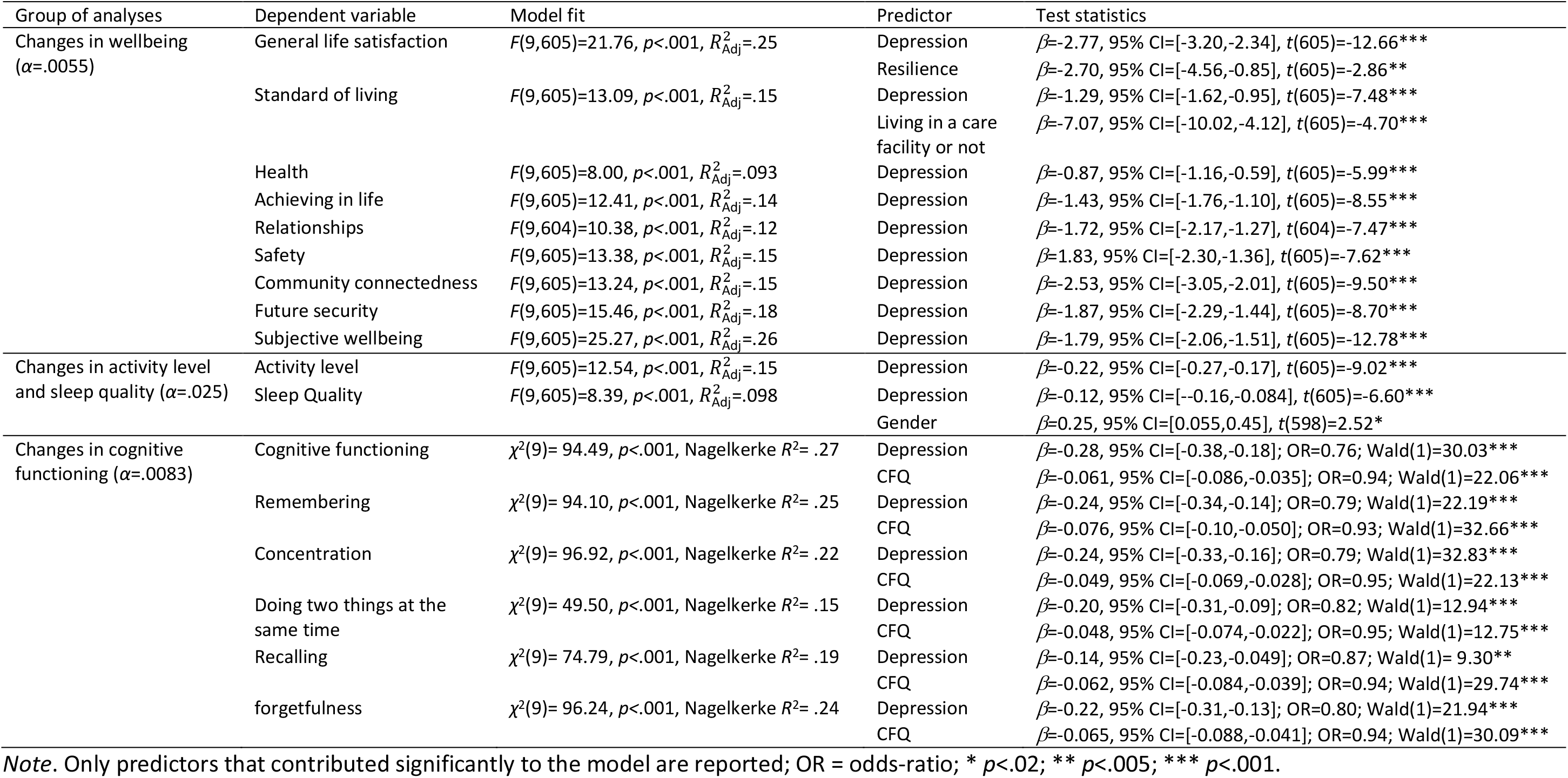
Statistical results of the 17 linear and ordinal logistic regression analyses conducted for changes in wellbeing, activity level and sleep quality (all based on the calculated difference scores) and cognitive functioning (based on the subjective cognitive change questions) with gender, age, whether the participant lived alone or not, whether the participant lived in a care facility or not, monthly individual net income, depression (i.e., GDS-15 total score), social network (i.e., LSNS-6 total score), general susceptibility to cognitive failures (i.e., CFQ total score) and resilience (i.e. BRS total score) as predictors. Both the fit of the full model (Model fit) and the test statistics for the significant predictors are reported.

*Changes in wellbeing* (α=.0055). With regards to general life satisfaction, the model indicated that at least one predictor significantly contributed to explaining changes in this wellbeing item. Changes in general life satisfaction showed a significant negative linear relation with depression. This implies that an increase in depression was associated with a (stronger) decrease in satisfaction with standard of living during the COVID-19 period. Changes in general life satisfaction also showed a significant negative linear relation with resilience. However, as this association is contrary to the positive correlation coefficient observed between these two variables (r=.11, see Supplementary Table 2), this could rather indicate a suppressor situation (see Darlington, 1968; Paulhus, Robins, Trzesniewski, & Tracy, 2004), leading us to be cautious about this effect. With regards to satisfaction with standard of living, the model indicated that at least one predictor significantly contributed to explaining changes in this wellbeing item. Changes in satisfaction with standard of living showed a significant negative linear relation with both depression and whether the participant lived in a care facility or not. This implies that an increase in depression and living in a care facility as compared to not living in a care facility (as “not living in a care facility” was set as the reference category of our dummy variable) were associated with a (stronger) decrease in satisfaction with standard of living during the COVID-19 period. For all other PWI-A items and total score, models always indicated that at least one predictor significantly contributed to explaining changes in these items or total score. For all these models, there was only a significant negative linear relation between depression and The impact of the COVID-19 pandemic on an aging population changes in these wellbeing items and total score. This implies that an increase in depression was associated with a decrease in satisfaction with all domains and in subjective wellbeing during COVID-19. None of the other predictors contributed significantly to the models (all p>.007).

*Changes in activity and sleep* (α=.025). The model indicated that at least one predictor significantly contributed to explaining changes in activity level. There was a significant negative linear relation between depression and changes in activity. This implies that an increase in depression was associated with a decrease in activity level during the COVID-19 period. None of the other predictors contributed significantly to the model (all p≥. 12). With regards to *sleep quality*, the model indicated that at least one predictor significantly contributed to explaining changes in sleep quality. There was a significant negative linear relation between depression and changes in sleep quality and a positive relation between gender and changes in sleep quality. This implies that an increase in depression and being male as compared to female (as “male” was set as the reference category of our dummy variable) were associated with a (stronger) decrease in sleep quality during COVID-19. None of the other predictors contributed significantly to the model (all p>.12).

### Changes in cognitive functioning

For all six subjective cognitive change questions, the ordinal logistic models had a significantly better fit compared to the null model. In all models, two predictors always contributed significantly to explaining the changes in cognitive functioning. When depression increased with 1 unit, the ordered log-odds of being in a higher category of the response scale of the subjective cognitive change questions (with a higher category indicating fewer problems, see Methods section) decreased by 0.28 for general cognitive functioning and by 0.24, 0.24, 0.20, 0.14 and 0.22 for remembering, concentration, doing two things at the same time, recalling and forgetfulness, respectively, while the other variables in the model are held constant. When the susceptibility to cognitive failures (i.e., CFQ total score) increased with 1 unit, the ordered log-odds of being in a higher category of the response scale of the subjective cognitive change questions decreased by 0.061 for general cognitive functioning and by 0.076, 0.049, 0.048, 0.061 and 0.065 for remembering, concentration, doing two things at the same time, recalling and forgetfulness, while the other variables in the model are held constant. None of the other predictors contributed significantly to the models (all *p>*.015).

We note that including the four variables describing the number of contacts participants had had during the past week (see Supplementary Table 1) as additional predictors in the models, did not lead to significant effects (all *p*≥.045) and did not change the pattern of results, except that the association between resilience and changes in general life satisfaction was no longer significant with the set *α* (*p*=.008). For reasons of parsimony we therefore opted not to include these additional predictors.

## DISCUSSION

The COVID-19 pandemic had an enormous impact on older adults aged 65 years or older. The risk of social isolation and loneliness due to governmental regulations raises concerns about the mental health and cognitive functioning of this population. Based on recent editorials about the impact of the COVID-19 pandemic on older adults and literature on the impact of disasters, we hypothesised that wellbeing, level of activity, quality of sleep and cognitive functioning would be severely impacted. To address this, we assessed how older adults are currently doing. Using an online survey with self-report measures, we studied the impact of the COVID-19 period on wellbeing, level of activity, quality of sleep and cognitive functioning of a general population of older adults aged 65 years or older. In addition, vulnerability and protective factors were examined that might have influenced the impact of the COVID-19 period.

In line with our hypothesis, it became clear that the COVID-19 period had a significant impact on older adults. We observed a significant decrease in wellbeing, activity level and sleep quality during the COVID-19 period as compared to before COVID-19. This is not in line with the first studies being published on how older adults responded to the pandemic, which did not observe a severe impact on older adults’ wellbeing (Kivi et al., in press; Lopez et al., in press; van Tilburg et al., in press). But as these studies were completed during the early onset of the COVID-19 pandemic, this might highlight that the severity of the impact increased when the pandemic (and the imposed restrictions) further progressed. Most older adults indicated that their cognitive functioning had not changed during the COVID-19 period, although a subgroup of 8% indicated a decrease.

When examining possible vulnerability and protective factors, reported changes in wellbeing, activity level, sleep quality and cognitive functioning were especially related to depression. These findings suggest that depression might be a vulnerability factor that influenced the impact of the COVID-19 period on older adults. However, given the cross-sectional nature of our data, it is not clear if depression was a precursor, already present before the COVID-19 period, or if depression was triggered and/or intensified during the COVID-19 period. Previous studies showed that depression in itself is negatively correlated with wellbeing, activity and sleep (Maglione et al., 2012; Malone & Wachholtz, 2017; Roshanaei-Moghaddam, Katon, & Russo, 2009). However, the socioemotional selectivity theory (Carstensen, 1995) could also explain why depression increased as a consequence of the COVID-19 period. This theory proposes that older adults use their social network as a buffer against negative experiences (Swift et al., 2014). As the COVID-19 pandemic led to a decrease of older adults’ social network and contacts, this emotional buffer might have disappeared, which in turn could have paved the way for depression. In order to shed more light on the exact role of depression in the observed COVID-19 changes, we aim to longitudinally follow a substantial subsample of these older adults to study how their reported changes in wellbeing, activity level, sleep quality and cognitive functioning, and the role of depression therein, evolve with the fluctuating COVID-19 situation over time.

Next to depression, other variables such as living in a care facility or not and gender were related to changes in one specific domain of wellbeing and sleep quality, respectively. Susceptibility to cognitive failures was related to changes in cognitive functioning during the COVID-19 period.

Some limitations of this study need to be addressed. First of all, the sample of older adults in this study is rather homogenous in some respects. Participants were all Dutch-speaking older adults living in Flanders. Moreover, most were in good health and their socioeconomic status was high, as indexed by educational level and income. Therefore, the results of this study might be difficult to generalise to the whole population of older adults and additional studies in different countries, and more heterogeneous samples of older adults are needed. Moreover, since this study was administered online, a part of this older adult population could probably not be reached. Third, the design of this study was cross-sectional and administered during the COVID-19 period. Questions regarding the period before COVID-19 were therefore necessarily retrospective, which might lead to biased self-reports (Sato & Kawahara, 2011). In addition, this study relied on subjective self-report which can differ from objective states (e.g. Burmester, Leathem, & Merrick, 2016). For example, it has already been shown that cognitive ability tends to be overestimated with increasing age (van der Ham, van der Kuil, & Claessen, 2019). During the ongoing COVID-19 pandemic, experimental studies administered to a sample of older adults repeatedly over time (e.g., when the pandemic activity decreases or flares up again in new waves) could shed more light on the actual effect of the COVID-19 period on cognitive functioning.

To summarize, the COVID-19 pandemic had a severe impact on the wellbeing, activity level and sleep quality of older adults. Only a small group of participants reported a decline in cognitive functioning. All changes reported during the COVID-19 period were strongly related to depression. Findings of this study are important because they give a first, thorough assessment of the mental health of older adults during this COVID-19 pandemic. This study showed that the concerns raised about the wellbeing of older adults are justified, and that this group at risk requires the attention of governments and healthcare. Furthermore, this study exposed that when we are faced with extreme stressors, such as COVID-19, in the future, prevention and intervention strategies are needed to aid older adults to prepare for and cope with them, especially for those at risk of depression (see also suggestions by Armitage & Nellums, 2020; Tian et al., 2020). As COVID-19 has complicated the accessibility of this population through social distancing, nursing home restrictions, and an overwhelmed health system, we are urged to explore novel ways to reach older adults (Chong, Curran, Ames, Lautenschlager, & Castle, 2020).

## METHODS

### Participants

Participants were recruited through social media, radio news, targeted newsletters and discussion fora and electronic mailings to all directors of assisted living facilities and nursing homes in Flanders. Only adults aged 65 years or older with a thorough knowledge of Dutch and living in Belgium could participate in the study. Only data of participants who filled in at least 50% of the survey were retained for analysis to ensure sufficient data quality. If a participant completed the survey more than once (N= 5), only the most complete, or, in case all entries were equally complete, the first entry was retained for analyses. In total, 640 participants meeting these criteria took part in the survey study.

We have complied with all relevant ethical regulations. All participants provided written informed consent. This study was approved by the Social and Societal Ethics Committee (SMEC) from KU Leuven (G-2020-1987). For their participation, participants could win one of 16 gift certificates via a random draft.

### Material

The online survey was created and data were generated using Qualtrics software (Qualtrics, Provo, UT). The language of the survey was Dutch. The survey existed of several general and demographic questions and questionnaires.

#### General and demographic questions

We asked participants several general questions assessing their year of birth, gender, country of residence, nationality, postal code, living situation, educational level, current and previous work situation, monthly individual net income, age-related diseases and whether the participant and/or any of their close relatives or friends had been infected with the corona virus.

#### Subjective cognitive functioning

To assess subjective cognitive functioning, the Dutch version of the *Cognitive Failures Questionnaire* (CFQ; Broadbent, Cooper, Fitzgerald, & Parkes, 1982;The impact of the COVID-19 pandemic on an aging population Merckelbach, Muris, Nijman, & de Jong, 1996) was used. The CFQ exists of 25 items assessing self-reported frequency of failures in several cognitive domains (e.g., perception, attention, memory and action). In our study, participants indicated how often these failures occurred during the past month (i.e., during the COVID-19 period) on a 5-point scale ranging from “very often” (=4) to “never” (=0). We added a response option “not applicable”, since some situations described in the questionnaire might not have occurred during the COVID-19 period for some participants (e.g., “Do you fail to see what you want in a supermarket (although it’s there)?”). A total score across all items, varying between 0 and 100, provides a measure of the general susceptibility to cognitive failures with a higher score indicating a higher susceptibility. The “not applicable” scores were not included to calculate the total score. For participants who indicated “not applicable” on more than 50% of the items (N=15), no CFQ total score was computed. The Dutch version of the CFQ has been validated across the adult life span, including older adults (Rast, Zimprich, Van Boxtel, & Jolles, 2009). The internal consistency of the CFQ in the current study was high with Cronbach’s *α*=.92.

#### Subjective cognitive change

Additionally, we assessed whether participants felt that their general cognitive functioning had changed during the COVID-19 period using a 3-point scale: Yes, it has decreased (=1); No, it has not changed (=2); Yes, it has improved (=3). We also asked them whether they had experienced the following cognitive problems during the COVID-19 period: problems to remember things, to concentrate on something, to do two things at the same time, to recall things and forgetfulness. They indicated the frequency of these problems in comparison to the period before the COVID-19 period on a 5-point scale with labels “a lot more than before” (=1), “more than before” (=2), “not more or less than before” (=3), “less than before” (=4), “a lot less than before” (=5). The internal consistency of these six subjective cognitive change questions was high with Cronbach’s *α*=.86.

#### Depression

To assess self-reported depression, the *Geriatric Depression Scale-15* (GDS-15; Sheikh & Yesavage, 1986; Bleeker, Frohn de Winter, & Cornelissen, 1985) was used. This scale exists of 15 Yes/No items and is designed to assess depressive symptoms in older populations. Using a The impact of the COVID-19 pandemic on an aging population score key, each item receives a score of 0 or 1 and these scores are then summed leading to a total score between 0 and 15, with higher scores indicating more depressive symptoms. The psychometric qualities of the GDS-15 have been supported by numerous studies, also in Dutch older populations (e.g., de Craen, Heeren, & Gussekloo, 2003). The internal consistency of the GDS-15 in the current study was high with Cronbach’s α=.81.

#### Activity and sleep

Participants were asked to evaluate their activity level and quality of sleep both before the COVID-19 period (retrospectively), and during the past week (i.e., during the COVID-19 period). These four questions were answered on an 11-point scale ranging from 0 (i.e., “not active at all” and “very poor quality of sleep” respectively) to 10 (i.e., “extremely active” and “very good quality of sleep” respectively).

#### Wellbeing

Wellbeing was assessed using the Dutch version of the *Personal Wellbeing Index – Adults* (PWI-A; International Wellbeing Group, 2013; Van Beuningen & de Jonge, 2011). The PWI-A is a multidimensional scale measuring life satisfaction in seven different domains (standard of living, health, achieving in life, relationships, safety, community connectedness and future security). An additional item assessing general life satisfaction can also be administered. All eight items are scored on an 11-point scale ranging from “no satisfaction at all” (=0) to “completely satisfied” (=10). The raw scores are converted to a 0-100 scale, with higher scores indicating more satisfaction. The scores for each domain can be studied individually, and the seven domain items (excluding the general life satisfaction item) can be summed to provide an index of “subjective wellbeing”. The psychometric properties of the PWI-A have been reported as satisfactory for older adults (Forjaz et al., 2011) and for a Dutch sample (Van Beuningen & de Jonge, 2011). In the current study, participants filled in the PWI-A twice: they indicated their satisfaction for each domain once (retrospectively) for the period before COVID-19 and once for the past month (i.e., during the COVID-19 period). The internal consistency in the current study was high for the PWI-A total score (i.e., 7 items) relating to the pre-COVID-19 period (Cronbach’s α=.91) and to the past month (Cronbach’s α=.91).*Social network*. The *Lubben Social Network Scale-6* (LSNS-6; Lubben et al., 2006), specifically constructed for older adults, was used to assess social network, support and isolation. It consists of six items: three items evaluating family ties and three comparable items evaluating non-family ties. These items are scored on a 6-point scale where participants indicate the number of ties (i.e., 0, 1, 2, 3 or 4, 5 to 8, 9 or more). A sum score can be calculated, ranging between 0 and 30, with a higher score indicating more social engagement. The psychometric properties of the LSNS-6 are good (Lubben et al., 2006), although we were unable to obtain data for the Dutch version specifically. The internal consistency of the LSNS-6 in the current study was high with Cronbach’s α=.82. Next to the LSNS-6, we also asked participants how many contacts they had (not taking into account cohabitants) during the past week in real life outside, in real life inside, by telephone and via the internet (e.g., skype, whatsapp), using the same response scale of the LSNS-6.

#### Resilience

Resilience, or the ability to bounce back or recover from stress, was assessed using the Dutch version of the *Brief Resilience Scale* (BRS; Smith et al.; 2008; Soer, Bieleman, & Schreurs, 2019). The BRS exists of six items measuring the degree of individual resilience. The items are scored on a 5-point scale ranging from “strongly disagree” to “strongly agree”. After reversing items 2, 4 and 6, a mean score is calculated which ranges between 1 and 5, with a higher score indicating more resilience. The validity of the BRS has been confirmed in older adults (da Silva-Sauer et al., 2020) and for the Dutch version (Soer et al., 2019). The internal consistency of the BRS in the current study was high with Cronbach’s α=.81.

### Procedure

The survey existed of a first page providing participants with information about the study and an informed consent form. After participants had read the instructions and informed consent, they were asked to explicitly check a box if they agreed to participate and to confirm that they were 65 years or older.

Afterwards, participants first completed the general and demographics questions. Next, the questionnaires were administered in the following order: CFQ, subjective cognitive change questions, GDS-15, questions regarding activity level and sleep quality, PWI-A, additional social network question, LSNS-6 and BRS. Finally, participants could provide their contact details to contact them for follow-up surveys or in case they had won a gift certificate. After completing the survey, we provided participants with web links and a telephone number where they could obtain more information about COVID-19 and psychological care if needed. The median number of minutes it took participants to complete the survey was 20. The survey was launched on May 19 2020 and closed on June 22 2020.

### Statistical analyses

First, we summarized the participant characteristics and the Pearson correlations between the main constructs. We considered correlations as small, medium or strong when the correlation coefficient was below .30, between .30 and .50 and above .50, respectively (based on Cohen, 1988).

Second, we compared participants’ report of their current wellbeing, activity level, sleep quality and cognitive functioning (i.e., *during* the COVID-19 period) with their retrospective report of these variables *before* COVID-19. We calculated difference scores between the reports during and before the COVID-19 period for activity level, sleep quality and the PWI-A items and total score (i.e., report_during_ – report_before_, where negative scores indicate a decrease in the report during the COVID-19 period compared to before). From now on, we will refer to these as the *difference scores*. We reported the descriptive statistics for the difference scores and the subjective cognitive change questions. Furthermore, we conducted two-sided paired-samples t-tests with Bonferroni correction to compare the reports during and before the COVID-19 period. As 11 comparisons were conducted (i.e., for the eight PWI-A items and total score, activity level and sleep quality). Cohen’s *d* (calculated as the difference between the means divided by the standard deviation) were reported. We The impact of the COVID-19 pandemic on an aging population considered effect sizes as small, medium or strong when the Cohen’s *d* was around .20, .50 and .80, respectively (based on Cohen, 1988).

Finally, we conducted linear or ordinal logistic regression analyses to determine the contribution of potential vulnerability and protective factors, in explaining changes in wellbeing, activity level and sleep quality (i.e., the 11 difference scores) and cognitive functioning (i.e., the six subjective cognitive change questions). This led to a total of 17 dependent variables and hence 17 separate regression analyses. These analyses were grouped in three parts focused on changes in wellbeing (i.e., nine analyses), changes in activity and sleep (two analyses) and changes in cognitive functioning (i.e., six analyses). Bonferroni correction was applied for each group of analyses, leading to *α* being set at .0055 for wellbeing, .025 for activity and sleep and.0083 for cognitive functioning. For changes in wellbeing, activity level and sleep quality, linear regression analyses were performed. Given the ordinal nature of the subjective cognitive change questions, we conducted ordinal linear regression analyses for changes in cognitive functioning. The following variables were included in each regression analysis as predictors: gender, age, whether the participant lived alone or not (derived based on the reported number of cohabitants, see Supplementary Table 1), whether the participant lived in a care facility or not (derived based on the reported living situation, see Supplementary Table 1), the reported monthly individual net income (to index socio-economic status), depression (i.e., GDS-15 total score), social network (i.e., LSNS-6 total score), general susceptibility to cognitive failures (i.e., CFQ total score) and resilience (i.e. BRS total score). Categorical predictors were dummy recoded. The presence of multicolinearity was examined using a cut-off of <5 for the Variance Inflation Factor (VIF; Sheather, 2009).

## Data Availability

The anonymized, raw data that support the findings of this study are available in the Open Science Framework (OSF; https://osf.io/re7sm/) with the identifier doi: 10.17605/OSF.IO/RE7SM.

## DATA AVAILABILITY STATEMENT

The anonymized, raw data that support the findings of this study are available in the Open Science Framework (OSF, https://osf.io/re7sm/) with the identifier “doi: 10.17605/OSF.IO/RE7SM”.

## CODE AVAILABILITY

The SPSS code used to generate results that are reported in the paper is available upon request.

## ACKNOWLEDGMENTS

The authors wish to thank all participants for completing the survey and all nursing staff who helped participants to complete the survey.

## AUTHOR CONTRIBUTIONS

S.D.P., C.G. and E.V.D.B. designed the study; S.D.P. and E.V.D.B. created the survey; S.D.P., C.G. and E.V.D.B. recruited the participants; S.D.P. and E.V.D.B. analyzed the data and wrote the manuscript; C.G., E.D., M.-A.V and R.D.R provided feedback on previous drafts of the manuscript, and provided suggestions for the analysis strategy.

## COMPETING INTERESTS STATEMENT

No competing interests exist.Means (standard deviations) and Pearson correlations between age and the total scores of the questionnaires in the survey.

## Notes

### Competing Interest Statement

The authors have declared no competing interest.

### Funding Statement

No external funding was received for this study.

### Author Declarations

This study was approved by the Social and Societal Ethics Committee (SMEC) from KU Leuven (G-2020-1987).

